# Predicting HIV Drug Resistance among Persons Living with HIV/AIDS in Sub-Saharan Africa using Population-based HIV Impact Assessment Surveys: 2015-2019

**DOI:** 10.1101/2024.04.11.24305688

**Authors:** Edson Nsonga, Mtumbi Goma, Wingston Felix Ng’ambi, Cosmas Zyambo

**Author notes:** **Corresponding author information:** Edson Nsonga, University of Zambia, Department of Public Health and Family Medicine, Lusaka, Zambia.

## Abstract

**Introduction:** HIV drug resistance (HIVDR) remains a significant challenge in sub-Saharan Africa (SSA), where access to effective treatment and healthcare resources varies widely. Socioeconomic status, demographic factors, clinical parameters, and regional disparities have been associated with patterns of HIVDR across SSA. Understanding the interplay of these factors is crucial for designing effective interventions to mitigate the impact of HIVDR and improve treatment outcomes in the region.

**Methods:** We conducted a secondary analysis of the Population-based HIV Impact Assessment (PHIA) HIV drug resistance datasets from Cameroon, Malawi, Eswatini, Ethiopia, Namibia, Rwanda, Tanzania, Zambia and Zimbabwe. All recipients of care aged between 15+ years were included in this analysis. The outcome of interest was whether a person had HIVDR resistant strains or no HIVDR resistant strains. Predictive analysis, chi-square test, univariable and multivariable logistic regression analyses were conducted in R. Statistical significance was set at P<0.05.

**Results:** The total sample size across the nine countries was 1008. Tanzania had the highest representation (16.8%), followed by Zambia (16.3%) and Zimbabwe (14.2% while Rwanda had the lowest representation (5.1%). Significant associations were observed between ARV status, viral suppression, country of residence and HIVDR in SSA. Individuals residing in Rwanda had significantly higher odds of HIVDR (adjusted OR = 3.63, 95% CI: 1.22-11.0, p = 0.021) compared to other countries. Additionally, individuals with suppressed viral loads had significantly lower odds of HIVDR (adjusted OR = 0.31, 95% CI: 0.21-0.45, p < 0.001), while those on ART exhibited higher odds of HIVDR (adjusted OR = 2.6, 95% CI: 1.75-3.91, p < 0.001).

**Conclusion:** This study focused on how clinical and sociodemographic factors influence HIVDR patterns in SSA. To mitigate the effects of HIVDR and improve treatment outcomes in the region, it is critical to address barriers to treatment access and adherence and upgrade the healthcare system.

## INTRODUCTION

In the past ten years, there has been an unparalleled rise in the use of antiretroviral therapy (ART), saving the lives of tens of millions of individuals living with HIV/AIDS globally. By the end of 2021, approximately 28.7 million out of an estimated 38.4 million people living with HIV were receiving ART worldwide(1). Despite the progress in expanding access to ART, the challenge of HIV drug resistance has emerged. This phenomenon, caused by genetic mutations in the HIV virus, poses a serious threat to the effectiveness of antiretroviral drugs(2).

HIV drug resistance (HIVDR) is caused by one or more changes (mutations) in the genetic structure of HIV that affect the ability of a specific drug or combination of drugs to block replication of HIV. All current ARV drugs, including newer classes, are at risk of becoming partly or fully inactive because of the emergence of drug-resistant virus(1).

HIV drug resistance is classified into three categories. When people contract drug-resistant strains of HIV without having previously used antiretroviral drugs, this is known as transmitted HIV drug resistance (TDR). Acquired HIV drug resistance (ADR) develops during antiretroviral therapy (ART) because of drug selection pressure, in which antiretroviral drugs eliminate non-resistant HIV strains while allowing drug-resistant strains to thrive, causing treatment failure. Poor treatment adherence and ineffective drug regimens exacerbate the pressure. Pretreatment HIV drug resistance (PDR) is detected in people starting or restarting ART, resulting from TDR, ADR, or both, highlighting a complex interplay of factors that contribute to drug resistance development. PDR can be contracted through transmission from others or prior exposure to antiretroviral drugs, including those used for pre-exposure prophylaxis or to prevent mother-to-child transmission(3).

Four types of factors are associated with HIVDR especially for ADR: factors related to the program; factors related to the patient; factors related to the regimen and drugs; and factors related to the virus. When considering patient-related variables like socioeconomic status and demographics, it is important to remember that people frequently become weary of their ART because of its long-term side effects. This weariness is exacerbated by the demanding daily schedules, frequent doctor visits, and intricate drug plans. Additionally impeding factors are psychological ones, such as drug misuse, mental illness, and cognitive aging. In addition to making matters worse, socioeconomic issues like hunger, poverty, and underdevelopment can also make it more difficult for people to access medical care. Treatment adherence can also be impacted by demographic factors like age, gender, and location(3)

HIV drug resistance poses a significant challenge to effective HIV treatment and prevention efforts. Understanding the dynamics of HIVDR, including its prevalence, determinants, and spatiotemporal characteristics, is crucial for informing targeted interventions and public health strategies. A study was conducted to investigate HIVDR among people with HIV (PWH) in Florida from 2012 to 2017. This study aimed to estimate the prevalence of HIVDR, explore sociodemographic and socioecological determinants, and describe the spatiotemporal patterns of HIVDR in the region.

The HIVDR patterns observed in Florida were significantly shaped by socioeconomic factors. The discussion in the study emphasized the complex connection between socio-economic and demographic factors and patterns of HIV drug resistance (HIVDR) among individuals living with HIV in Florida. A key finding of the study was the strong link between lower socio-economic status (SES) indicators and increased rates of HIVDR. Countries with lower SES, as measured by unemployment and median household income, tended to have higher levels of HIVDR. This highlights the urgent need to address socio-economic disparities in HIV treatment and care in order to ensure fair access and outcomes for all populations(4).

Monitoring and treating HIV drug resistance trends is critical to the highly effective use of highly active antiretroviral therapy (HAART) in the global fight against HIV/AIDS. A study conducted among Chinese HIV-infected patients receiving antiretroviral treatment (ART) revealed significant findings regarding HIV drug resistance (HIVDR). At baseline in 2009, the overall prevalence of HIVDR was 5.6%, with NNRTI mutations found in virtually all cases and three-fourths involving NRTI mutations. During the subsequent one-year follow-up, the incidence rate of HIVDR was 3.5 per 100 person years, with NNRTI mutations and NRTI mutations occurring at rates of 3.4 and 2.6 per 100 person years, respectively.

Several factors were identified as independently associated with HIVDR incidence. Patients who initiated treatment with didanosine-based regimens were 3.1 times more likely to develop HIVDR compared to those on lamivudine-based regimens. Additionally, individuals receiving care at rural township hospitals or village clinics had a 2.0-fold higher risk of HIVDR compared to those treated at country-level CDCs or hospitals. Notably, patients with baseline CD4 counts of 0–199 cells/µl were 2.3 times more likely to develop HIVDR than those with CD4 counts ≥350/µl, while those with baseline viral loads ≥1000 copies/ml had a 5.9-fold increased risk compared to those with viral loads <1000 copies/ml.

Furthermore, age and marital status were also significant factors associated with HIVDR incidence. The risk of developing HIVDR did not significantly differ across age groups (≤30, 31–40, 41–50, >50 years). However, marital status did not show a significant association with HIVDR incidence, with both married and unmarried individuals having similar risks.

The study’s discussion focused on how socioeconomic disparities and barriers to healthcare access worsen the higher incidence rates of HIVDR compared to some other countries, especially among patients on didanosine-based regimens and those receiving care at rural township hospitals or village clinics. Focusing on the significance of all-encompassing education and training programs for healthcare providers at every level, particularly in community-based settings, the discussion advocated for focused interventions to enhance treatment quality and adherence support(5).

In sub-Saharan Africa, where the burden of HIV/AIDS is particularly high, addressing HIV drug resistance is crucial to ensuring the long-term effectiveness of treatment programs. The region’s unique challenges, including limited healthcare resources, varying levels of access to care, and diverse cultural and socioeconomic factors, contribute to the complexity of predicting and mitigating drug resistance.

Studies specific to sub-Saharan Africa have identified several contextual factors that influence the emergence of HIV drug resistance. Socioeconomic status is implicated to be a determinant, with lower-income individuals facing barriers to consistent treatment access and adherence. Geographical disparities, particularly between urban and rural areas, affect the availability of healthcare services and patient engagement in care(6).

It is crucial to recognize that sub-Saharan Africa encompasses diverse populations with varying degrees of vulnerability to HIV drug resistance. Key populations, including sex workers, men who have sex with men, and transgender individuals, may face unique challenges that impact their access to and engagement in care(7). Gender disparities, prevalent in many sub-Saharan African countries, can also influence treatment outcomes and adherence patterns(8).

In study conducted by Moyo (9) on comprehensive analysis of HIV drug resistance (HIVDR) among HIV-positive individuals in South Africa using data from the 2017 national HIV household survey. Among the key findings, HIVDR was detected in 27.4% of virally unsuppressed respondents, with the most prevalent resistance observed to non-nucleoside reverse transcriptase inhibitors (NNRTIs). Notably, 55.7% of individuals on antiretroviral therapy (ART) had HIVDR, highlighting the risk of transmission of resistant HIV. Resistance was particularly high among those who reported ARV use but tested ARV-negative (75.9%). The study also revealed low levels of resistance to second-line therapy, indicating appropriate limited prescription of these drugs. Additionally, there were no significant differences in HIVDR prevalence by age and sex. Overall, the findings underscored the importance of strengthening first-line ART regimens, enhancing treatment adherence strategies, and closely monitoring HIVDR to optimize HIV treatment outcomes and mitigate the risk of transmission of resistant strains.

Concerning rates of HIVDR coupled with virological failure (VF) were found in another study of HIV-positive adults in southern Mozambique at a district hospital outpatient clinic. Low-level viremia (LLV), overt VF (HIV-1 RNA levels ≥1000 copies/mL), and detectable viremia were present in a considerable fraction of the subjects. A significant portion of patients with VF possessed at least one HIVDR, with the bulk of them being resistant to both nucleoside and non-nucleoside RTIs (NRTIs and NNRTIs). Numerous factors, such as initiating antiretroviral therapy (ART) at WHO stage III/IV, being younger, having an estimated low adherence rate, being on ART for a longer period of time, and being illiterate, were linked to HIVDR and VF. According to the study, women made up a significant portion of the population and had a median age of 39. The level of education varied, but a significant portion only had an elementary education, had never gone to school, or were illiterate. Adherence problems were also apparent, as a significant portion of respondents indicated estimated low adherence and prior ART discontinuation (10).

Despite significant advancements in ART access in sub-Saharan Africa, there are still several obstacles to overcome. The complexity of treatment adherence is still a problem, with socioeconomic variables, the state of healthcare, stigma, and access to medical care all playing a role. Additionally, Sachathep et al (11) noted that the variability of HIV itself makes it difficult to forecast and handle medication resistance. To create successful treatments to lessen its effects, it is crucial to have a thorough understanding of the causes and predictors of HIVDR in this location. Therefore, this study was aimed at exploring the determinants of HIVDR among adult persons living with HIV from 10 countries in SSA using the PHIA data collected between 2015 and 2019.

## METHODS

### Study design

We conducted a secondary analysis of the PHIA data collected from SSA between 2015 and 2019. All adult persons living with HIV that had an assessment of HIV drug resistance were included in the analysis. Furthermore, only those aged between 15 and 80 years were included in this analysis. The PHIA utilized two-stage cluster design with census enumeration areas being the first stage to be sampled and the households being the second stage to be selected in order to achieve a representative sample (12) (13) (14). The samples are stratified by rural and urban location in all the countries. The numbers of households to be included in the surveys vary from country to country. For example, 30 households were selected per EA, with a minimum of 15 households in Zambia, Zimbabwe and Malawi while a maximum of 60 in Zimbabwe and Malawi and 50 in Zambia (12).

### Data management and analysis

We combined the adult and HIV DR PHIA data from Cameroon, Malawi, Eswatini, Ethiopia, Namibia, Rwanda, Tanzania, Zambia and Zimbabwe as prescribed in the PHIA Data use Manual (15). Data management was done in Stata (Stata Corp., Texas, USA). The following were the predictor variables: rural/urban location, survey year, socio-demographics (age, respondent’s sex (Male/Female), household wealth index, education level, marital status, ART status (on ART or not on ART), alcohol uptake, tobacco use (yes/no), alcohol use (yes/no), drug and substance use (yes/no), HIV viral suppression (yes/no)). The primary outcome variable was whether the APLHIV had HIV drug resistance or not.

We performed data analysis in Stata (Stata Corp., Texas, USA). We calculated counts, percentages, prevalence ratios (PR) and their associated 95% confidence intervals (95%CI). We included the socio-demographic and behavioural characteristics in bivariate analysis. We considered the variables for inclusion in the multivariate regression if they were significantly associated with the outcome at P<0.20 when doing bivariate analysis (16) (17). Both bivariate and multivariate prevalence ratios were calculated to determine the effects of each of the predictor variables on the binary endpoints of having HIV drug resistance strains. Data were presented using tables and figures wherever necessary.

### Ethical considerations

The initial ethics approval for the study protocols were obtained from the Centers for Disease Control and Prevention Institutional Review Board (IRB), the Columbia University Medical Center IRB, and relevant local regulatory bodies (18). In our case, we obtained permission to use this data from the International Center for AIDS Care and Treatment Programs (ICAP) at Columbia University. The PHIA datasets were accessed between 1^st^ June and 30^th^ June 2023 and downloaded from https://phia-data.icap.columbia.edu/datasets. As this study used secondary anonymised data, individual informed consent is not required.

## RESULTS

### Participant’s characteristics

**Characteristics of respondents across the nine sub-Saharan African countries.**

Overall, the analysis revealed that the majority of respondents fell within the age range of 35 and above, constituting 560 individuals, which accounted for 55.6% of the total respondents. In contrast, a slightly lower proportion of respondents, totaling 448 individuals, were aged between 15 and 34, representing 44.4% of the total respondents.

Examining the distribution by country, Tanzania exhibited the highest representation of younger individuals aged 15-34 years, with 78 individuals, constituting 46.2% of the total respondents, contrasting with Ethiopia, which had the lowest representation in this age group, with 32 individuals, comprising 35.2% of the total respondents. Conversely, Ethiopia presented the highest percentage of older respondents aged 35 and above, with 59 individuals, accounting for 64.8% of the total respondents, whereas Eswatini displayed the lowest, with 44 individuals, representing 37.9% of the total respondents.

Analysis by sex of the respondents revealed that a higher proportion of respondents were female, totaling 651 individuals, accounting for 64.6% of the total respondents, compared to male respondents, totaling 357 individuals, constituting 35.4% of the total respondents.

Across countries, Rwanda exhibited the highest proportion of male respondents, with 24 individuals, making up 47.1% of the total respondents, contrasting with Malawi’s lowest representation, with 37 individuals, constituting 30.3% of the total respondents. Conversely, Malawi reported the highest percentage of female respondents, with 85 individuals, accounting for 69.7% of the total respondents, whereas Eswatini presented the lowest, with 73 individuals, comprising 62.9% of the total respondents.

Analysis by ARV status revealed that a majority of respondents were on antiretroviral therapy (ART), totaling 720 individuals, which comprised 71.4% of the total respondents. In contrast, a smaller proportion of respondents, totaling 288 individuals, were not on ART, representing 28.6% of the total respondents.

Across countries, Eswatini demonstrated the highest proportion of individuals on ART, with 95 individuals, comprising 81.9% of the total respondents, whereas Tanzania exhibited the lowest, with 30 individuals, representing 17.8% of the total respondents. Conversely, Malawi indicated the highest percentage of individuals not on ART, with 24 individuals, making up 19.7% of the total respondents, whereas Tanzania reported the lowest, with 139 individuals, constituting 82.2% of the total respondents.

Analysis by harmful alcohol use revealed that the majority of respondents reported abstaining from harmful alcohol use, totaling 888 individuals, which accounted for 88.1% of the total respondents. In contrast, a smaller proportion of respondents, totaling 120 individuals, reported engaging in harmful alcohol use, representing 11.9% of the total respondents.

Across countries, Namibia exhibited the highest percentage of respondents reporting alcohol consumption, with 27 individuals, accounting for 27.8% of the total respondents, contrasting with Rwanda’s absence of harmful alcohol use among the respondents. Conversely, Rwanda displayed the highest percentage of respondents abstaining from harmful alcohol use, with 51 individuals, comprising 100% of the total respondents, while Namibia reported the lowest, with 70 individuals, representing 72.2% of the total respondents.

Analysis by residence revealed that a majority of respondents resided in urban areas, totaling 457 individuals, which comprised 45.3% of the total respondents. In contrast, a slightly lower proportion of respondents, totaling 551 individuals, resided in rural areas, representing 54.7% of the total respondents.

Across countries, Eswatini showcased the highest proportion of rural dwellers, with 92 individuals, making up 79.3% of the total respondents, whereas Zimbabwe displayed the lowest, with 43 individuals, representing 30.1% of the total respondents. Conversely, Malawi presented the highest percentage of urban residents, with 76 individuals, accounting for 62.9% of the total respondents, whereas Ethiopia reported the absence of rural respondents among its sample.

Analysis by level of education revealed that the majority of respondents had attained primary education, totaling 477 individuals, which accounted for 47.3% of the total respondents. In contrast, smaller proportions of respondents had attained secondary education (29.7%), tertiary education (12.1%), or had no formal education (10.9%).

Across countries, Ethiopia exhibited the highest percentage of respondents lacking formal education, with 17 individuals, comprising 18.7% of the total respondents, and Zimbabwe displaying the lowest, with 4 individuals, representing 2.8% of the total respondents. Conversely, Ethiopia reported the lowest percentage of respondents with tertiary education, with 4 individuals, making up 4.4% of the total respondents, while Cameroon presented the highest, with 55 individuals, accounting for 100% of the total respondents.

Analysis by wealth index revealed considerable heterogeneity in wealth distribution among respondents across the wealth index categories. The wealth index categories ranged from poorest (22.3%) to richest (17.8%).

Across countries, Namibia showcased the highest percentage of respondents classified as richest, with 16 individuals, representing 31.4% of the total respondents, in contrast to Malawi’s lowest representation, with 2 individuals, comprising 2.1% of the total respondents. Conversely, Malawi reported the highest percentage of respondents classified as poorest, with 38 individuals, making up 39.2% of the total respondents, while Namibia exhibited the lowest, with 24 individuals, accounting for 14.6% of the total respondents.

**Table 1:**
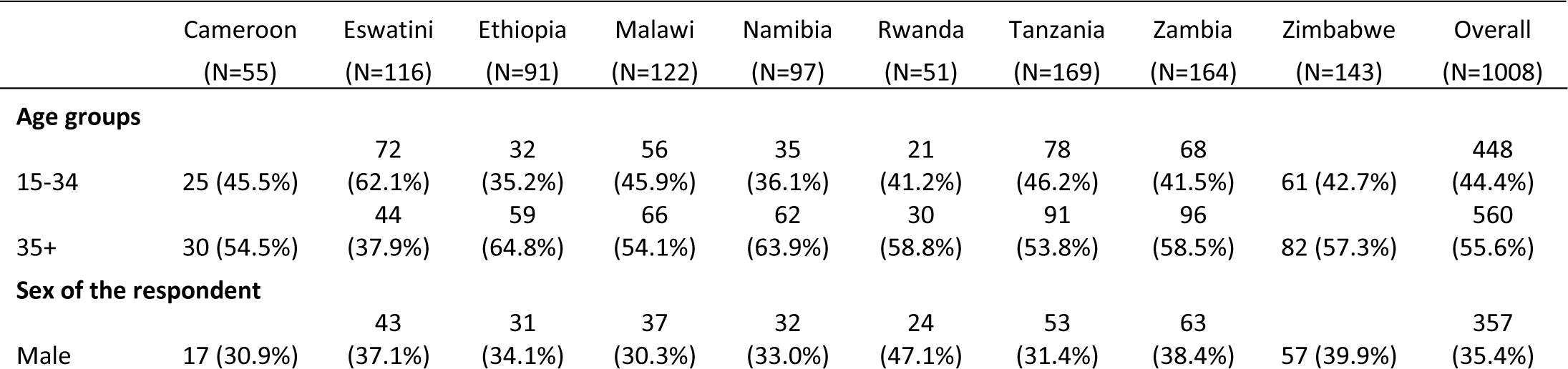
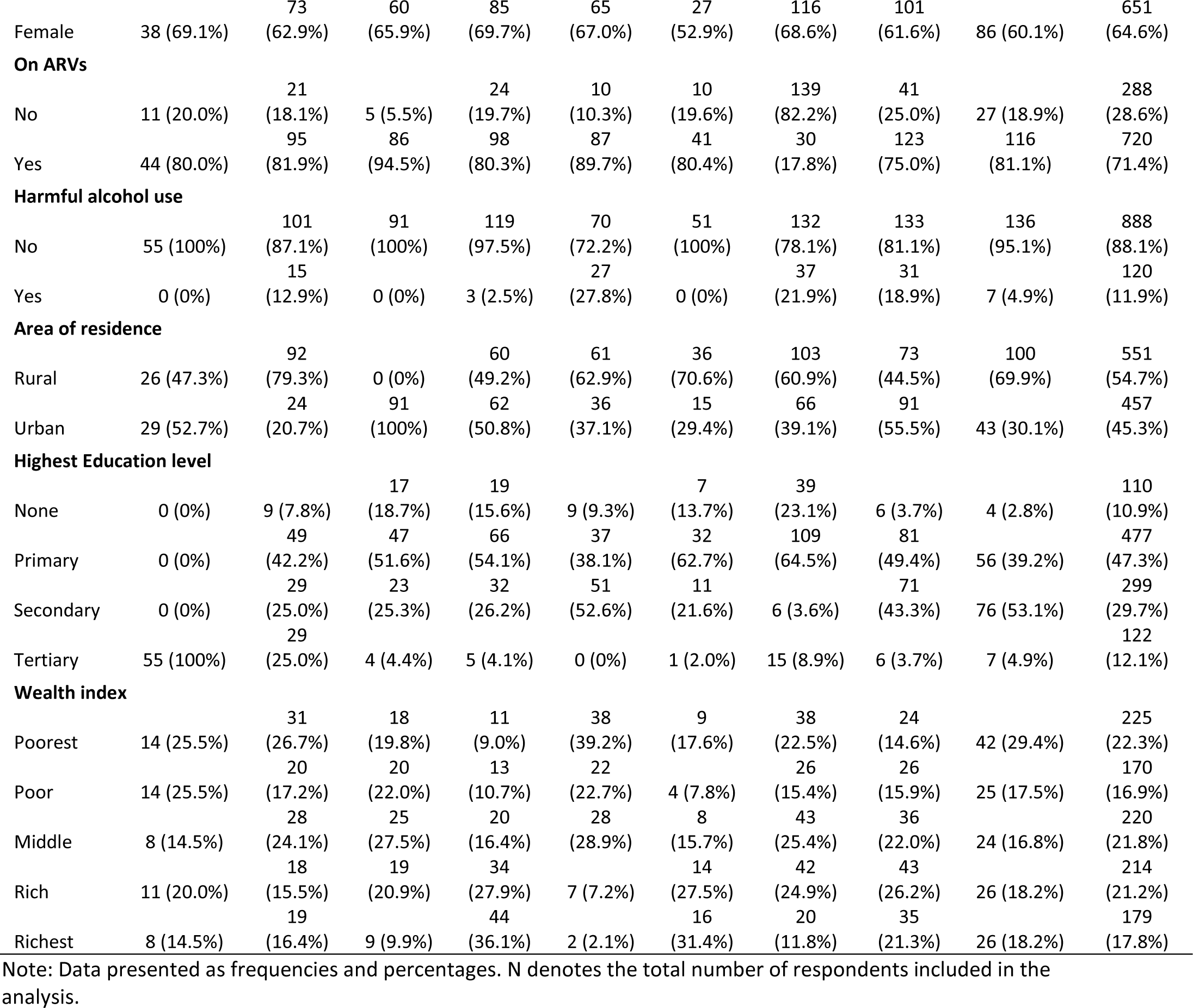
Characteristics of respondents in Rwanda, Namibia, Cameroon, Malawi, Eswatini, Zambia, Ethiopia, Zimbabwe and Tanzania between 2015 and 2019.

### Prevalence of HIV drug resistance

The prevalence of HIV drug resistance varied across the different countries surveyed. Rwanda had the highest prevalence of HIV drug resistance at 72.5%, followed by Namibia at 63.9% and Cameroon at 58.2%. Conversely, Eswatini and Zambia had relatively lower prevalence rates at 31.9% and 31.1%, respectively. Ethiopia and Zimbabwe demonstrated moderate prevalence rates at 25.3% and 23.1%, respectively. Tanzania had the lowest prevalence among the countries listed, with only 20.7% reporting HIV drug resistance.

**Figure 1:**
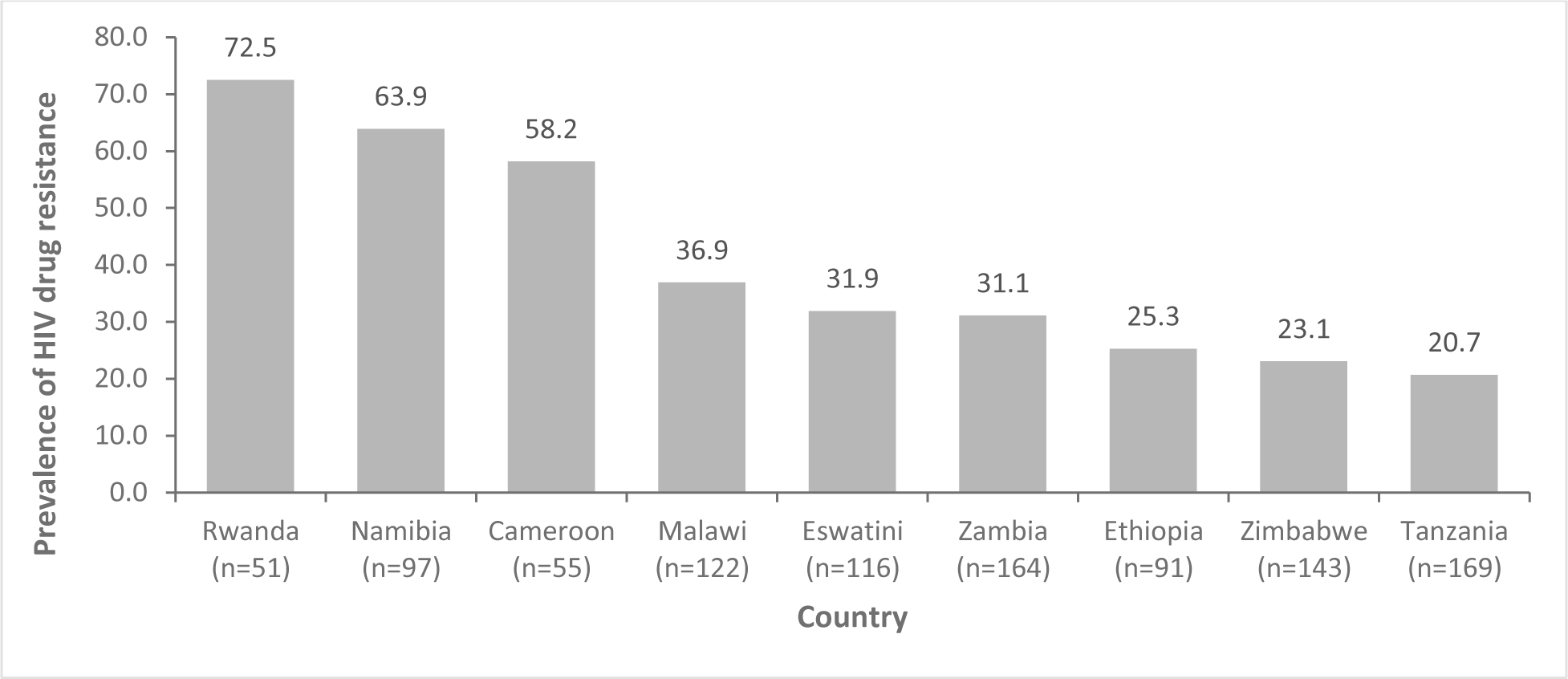
Prevalence of HIV drug resistance in Rwanda, Namibia, Cameroon, Malawi, Eswatini, Zambia, Ethiopia, Zimbabwe and Tanzania between 2015 and 2019.

### Correlates of HIV Drug Resistance

The chi-square analysis revealed significant associations between several variables and HIV drug resistance. Firstly, there was a notable disparity in HIV drug resistance across different countries (P < 0.001). Cameroon, Rwanda, and Zimbabwe exhibited relatively higher proportions of HIV drug resistance compared to other countries. However, no significant association was found between the sex of the respondent and HIV drug resistance (P = 0.8), indicating similar proportions of resistance among males and females.

Analysis by age groups, a significant association was observed (P = 0.018), with individuals aged 15-34 years showing a higher proportion of HIV drug resistance compared to those aged 35 years and above. Conversely, there was no significant association between the area of residence and HIV drug resistance (P = 0.15), indicating comparable resistance rates in both rural and urban areas.

Furthermore, a significant association was found between ARV usage and HIV drug resistance (P < 0.001). Individuals not on ARVs exhibited a higher proportion of HIV drug resistance compared to those on ARVs. However, marital status showed no statistically significant association with HIV drug resistance (P = 0.094), although single individuals demonstrated a slightly higher resistance proportion.

Similarly, no significant association was observed between wealth index and HIV drug resistance (P = 0.2), indicating similar resistance rates across different wealth categories. The highest education level also showed no significant association with HIV drug resistance (P = 0.3), suggesting comparable resistance proportions among individuals with different education levels.

However, a significant association was found between HIV viral load suppression and HIV drug resistance (P < 0.001). Individuals with unsuppressed viral loads exhibited a higher proportion of HIV drug resistance compared to those with suppressed viral loads. These findings underscore the complex interplay of various factors in influencing HIV drug resistance and highlight the importance of targeted interventions to mitigate its impact.

### Factors association with HIV drug resistance

The logistic regression analysis examined factors associated with HIV drug resistance, presenting both univariable and multivariable odds ratios (OR) along with their corresponding 95% confidence intervals (CI) and p-values.

The country of residence was significantly associated with HIV drug resistance. Individuals from countries like Eswatini, Ethiopia, Tanzania, Zambia, and Zimbabwe demonstrated significantly lower odds of HIV drug resistance compared to Cameroon (OR range: 0.19-0.47, 95% CI range: 0.10-0.65, P < 0.001).

Regarding the area of residence, individuals living in urban areas showed slightly lower odds of HIV drug resistance compared to those in rural areas in the univariable analysis (OR = 0.82, 95% CI: 0.63-1.07, P = 0.15). However, this association was not statistically significant in the multivariable analysis (OR = 0.98, 95% CI: 0.66-1.43, P = 0.9).

In the univariable analysis, no significant association was found between the sex of the respondent and HIV drug resistance (OR = 1.03, 95% CI: 0.79-1.36, p = 0.8). Similarly, after adjusting for other variables in the multivariable analysis, the association remained non-significant (OR = 1.01, 95% CI: 0.74-1.38, P > 0.9).

Although older age (35+) was associated with higher odds of HIV drug resistance compared to younger age (15–34) in the univariable analysis (OR = 1.37, 95% CI: 1.06-1.78, P = 0.018), this association became non-significant after adjustment in the multivariable analysis (OR = 1.3, 95% CI: 0.96-1.76, P = 0.1).

Being on antiretroviral therapy (ARVs) was significantly associated with higher odds of HIV drug resistance in both univariable and multivariable analyses (OR = 2.37, 95% CI: 1.74-3.27, P < 0.001 in univariable; OR = 2.6, 95% CI: 1.75-3.91, P < 0.001 in multivariable.

Individuals with HIV viral load suppression had significantly lower odds of HIV drug resistance compared to those without viral load suppression in both univariable (OR = 0.53, 95% CI: 0.39-0.72, P < 0.001) and multivariable (OR = 0.31, 95% CI: 0.21-0.45, P < 0.001) analyses.

No significant association was found between wealth index categories and HIV drug resistance in either univariable or multivariable analyses (OR range: 0.52-1.30, 95% CI range: 0.37-1.34, P > 0.05 for all wealth categories).

The association between harmful alcohol use and HIV drug resistance was not statistically significant in either univariable or multivariable analyses (OR range: 0.43-0.71, 95% CI range: 0.25-1.16, P > 0.05).

Factors associated with HIV drug resistance in Rwanda, Namibia, Cameroon, Malawi, Eswatini, Zambia, Ethiopia, Zimbabwe and Tanzania between 2015 and 2019.

| Characteristic | Univariable |  |  | Multivariable |  |  |
| --- | --- | --- | --- | --- | --- | --- |
|  | cOR | 95% CI | P-value | aOR | 95% CI | P-value |
| **Sex of the respondent** |  |  |  |  |  |  |
| Male | Ref |  |  | Ref |  |  |
| Female | 1.03 | 0.79, 1.36 | 0.8 | 1.01 | 0.74, 1.38 | >0.9 |
| **Area of residence** |  |  |  |  |  |  |
| Rural | Ref |  |  | Ref |  |  |
| Urban | 0.82 | 0.63, 1.07 | 0.15 | 0.98 | 0.66, 1.43 | 0.9 |
| **Highest Education level** |  |  |  |  |  |  |
| None | Ref |  |  | Ref |  |  |
| Primary | 1.04 | 0.68, 1.62 | 0.9 | 1.1 | 0.67, 1.82 | 0.7 |
| Secondary | 0.91 | 0.58, 1.45 | 0.7 | 0.83 | 0.47, 1.47 | 0.5 |
| Tertiary | 1.36 | 0.80, 2.33 | 0.3 | 1.17 | 0.54, 2.49 | 0.7 |
| **HIV viral load suppression** |  |  |  |  |  |  |
| No | Ref |  |  | Ref |  |  |
| Yes | 0.53 | 0.39, 0.72 | <0.001 | 0.31 | 0.21, 0.45 | <0.001 |
| **Age groups** |  |  |  |  |  |  |
| 15-34 | Ref |  |  | Ref |  |  |
| 35+ | 1.37 | 1.06, 1.78 | 0.018 | 1.3 | 0.96, 1.76 | 0.1 |
| **Country** |  |  |  |  |  |  |
| Cameroon | Ref |  |  | Ref |  |  |
| Eswatini | 0.34 | 0.17, 0.65 | 0.001 | 0.45 | 0.19, 1.03 | 0.063 |
| Ethiopia | 0.24 | 0.12, 0.49 | <0.001 | 0.47 | 0.17, 1.25 | 0.13 |
| Malawi | 0.42 | 0.22, 0.80 | 0.009 | 0.68 | 0.27, 1.71 | 0.4 |
| Namibia | 1.27 | 0.64, 2.51 | 0.5 | 1.65 | 0.63, 4.21 | 0.3 |
| Rwanda | 1.9 | 0.85, 4.37 | 0.12 | 3.63 | 1.22, 11.0 | 0.021 |
| Tanzania | 0.19 | 0.10, 0.36 | <0.001 | 0.34 | 0.13, 0.85 | 0.023 |
| Zambia | 0.32 | 0.17, 0.61 | <0.001 | 0.61 | 0.25, 1.50 | 0.3 |
| Zimbabwe | 0.22 | 0.11, 0.41 | <0.001 | 0.25 | 0.10, 0.62 | 0.003 |
| **Wealth index** |  |  |  |  |  |  |
| Poorest | Ref |  |  | Ref |  |  |
| Poor | 0.84 | 0.55, 1.27 | 0.4 | 0.84 | 0.52, 1.34 | 0.5 |
| Middle | 0.87 | 0.59, 1.28 | 0.5 | 1.03 | 0.66, 1.60 | >0.9 |
| Rich | 1.02 | 0.70, 1.50 | >0.9 | 1.3 | 0.79, 2.13 | 0.3 |
| Richest | 0.64 | 0.42, 0.98 | 0.041 | 0.66 | 0.37, 1.18 | 0.2 |
| **On ARVs** |  |  |  |  |  |  |
| No | Ref |  |  | Ref |  |  |
| Yes | 2.37 | 1.74, 3.27 | <0.001 | 2.6 | 1.75, 3.91 | <0.001 |
| **Harmful alcohol use** |  |  |  |  |  |  |
| No | Ref |  |  | Ref |  |  |
| Yes | 0.7 | 0.45, 1.05 | 0.094 | 0.71 | 0.43, 1.16 | 0.2 |

cOR= Crude Odds Ratio, aOR= Adjusted Odds Ratio, CI=Confidence Interval

### Predictive modelling of HIV drug resistance

We fitted the logistic regression on the training dataset which contained 80% of the dataset. The final model of the logistic regression (see Table 3) was used to determine the confusion matrix for the training dataset. The output of the logistic regression analysis using the test dataset is shown in Figure 3. Based on the predicted logistic regression model, the key determinants of HIVDR: HIV viral load, ARV status (On ARVs) and country.

**Figure 2:**
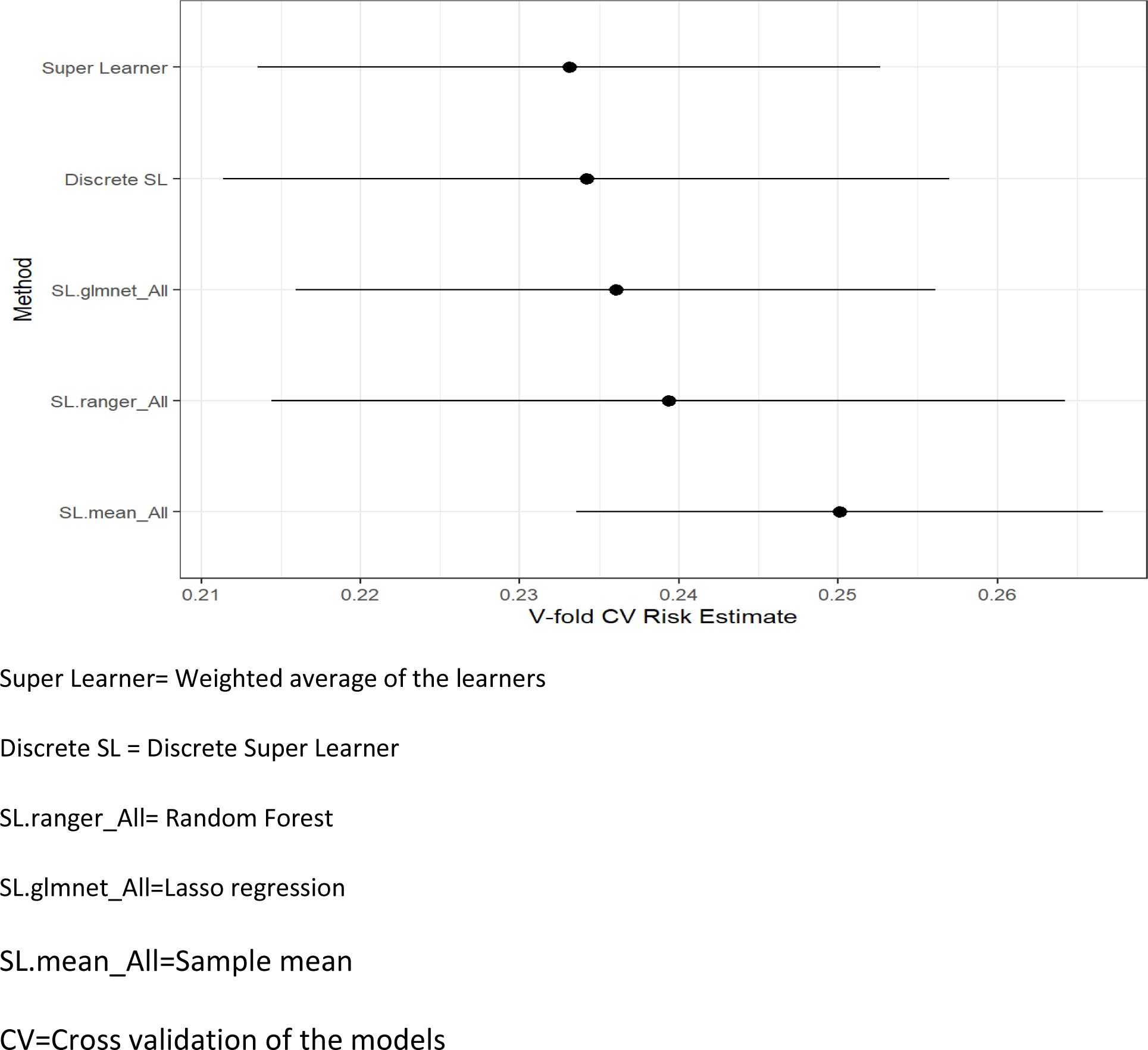
Plot the performance for the different supervised machine learning models fitted.

**Figure 3:**
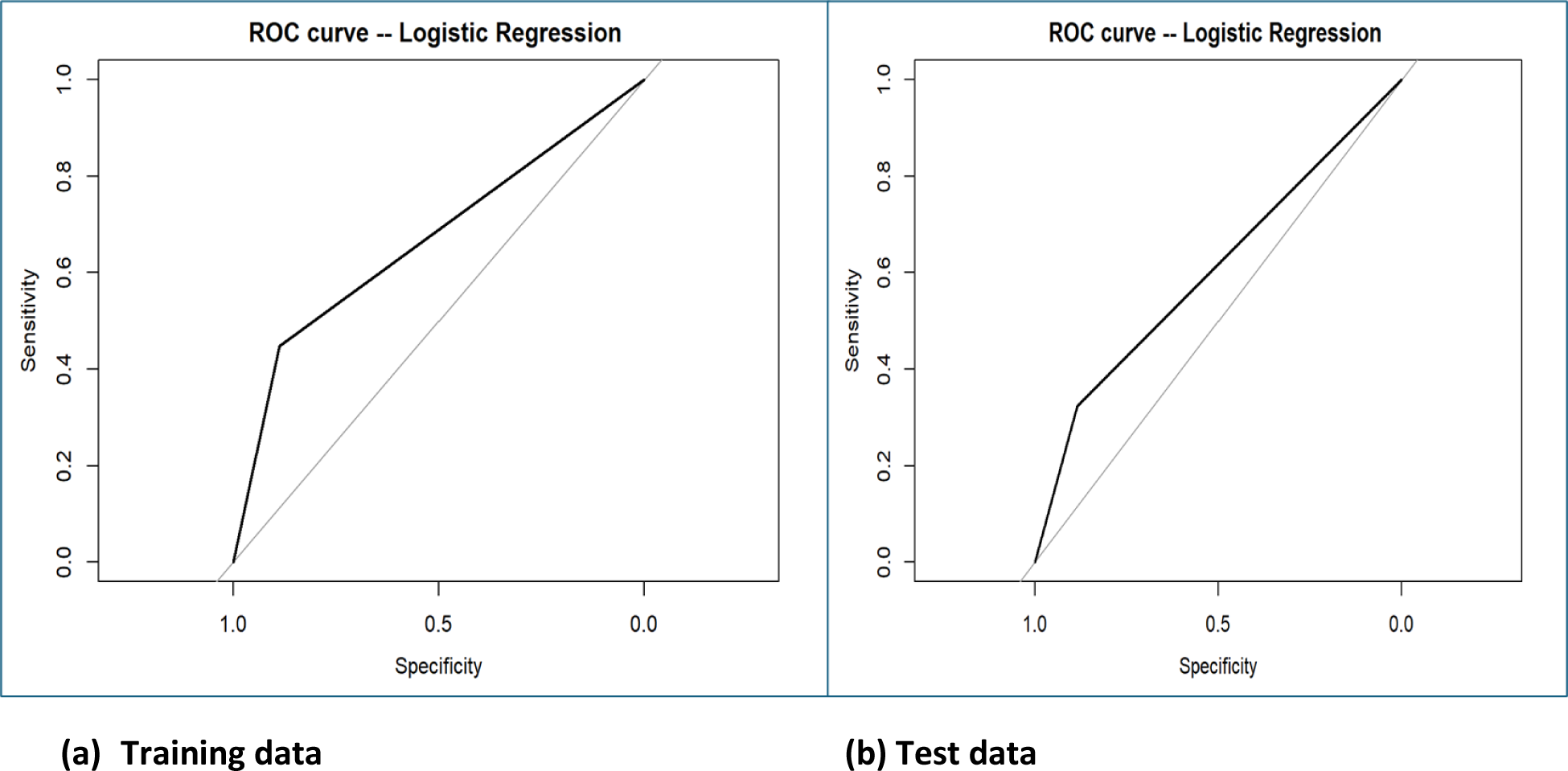
ROC curves for training and testing curves for screening HIV drug resistance in Rwanda, Namibia, Cameroon, Malawi, Eswatini, Zambia, Ethiopia, Zimbabwe and Tanzania between 2015 and 2019.

**Table 3:**
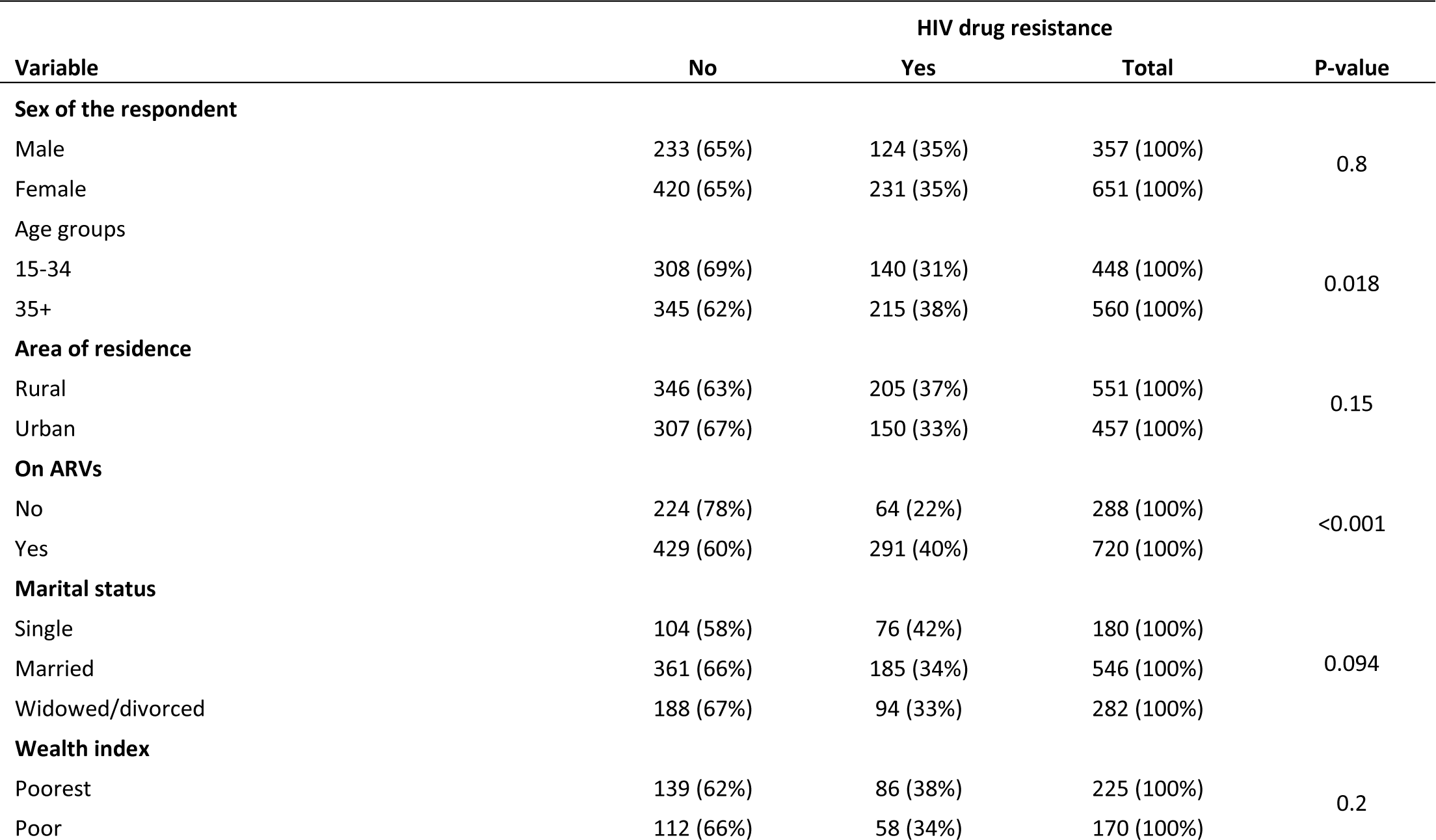
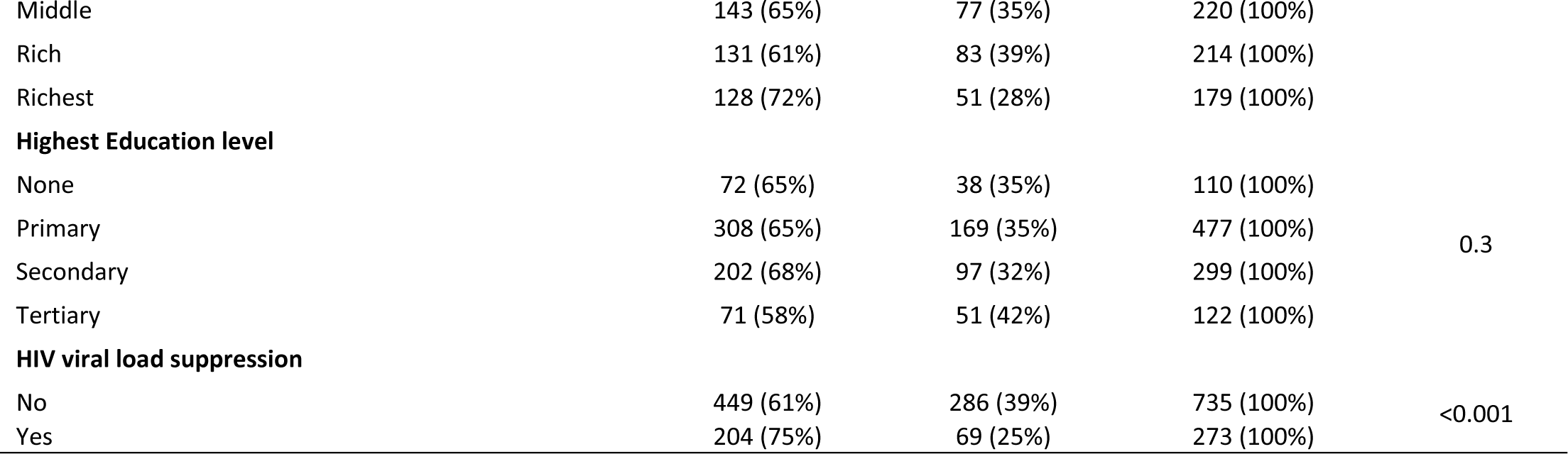
Correlates of HIV drug resistance in in Rwanda, Namibia, Cameroon, Malawi, Eswatini, Zambia, Ethiopia, Zimbabwe and Tanzania between 2015 and 2019.

### Receiver Operating Characteristic (ROC) Curves

In the presented ROC analysis, the model exhibited consistent accuracy in both the test and training datasets, each registering at 0.69. The precision values, representing the accuracy of positive predictions, were 0.61 for the test dataset and 0.68 for the training dataset.

**Table 4:** Confusion matrix for the accuracy of the fitted model of choice in classifying individuals for HIV drug resistance in Rwanda, Namibia, Cameroon, Malawi, Eswatini, Zambia, Ethiopia, Zimbabwe and Tanzania between 2015 and 2019.

| Training data |  |  |  | Test set |  |  |  |
| --- | --- | --- | --- | --- | --- | --- | --- |
| HIV drug resistance | No | Yes | TOTAL | HIV drug resistance | No | Yes | Total |
| No | **402** | 55 | 457 | No | **177** | 19 | 196 |
| Yes | 141 | **107** | 248 | Yes | 64 | **43** | 107 |
| Total | 543 | 162 | **705** | 0 | 241 | 62 | **303** |

**Table 5:** Performance metrics for predicting HIV drug resistance in Rwanda, Namibia, Cameroon, Malawi, Eswatini, Zambia, Ethiopia, Zimbabwe and Tanzania between 2015 and 2019.

| Parameter | Training dataset | Test Dataset |
| --- | --- | --- |
| Precision | 0.68 | 0.61 |
| Accuracy | 0.73 | 0.69 |
| Recall | 0.11 | 0.12 |
| False Positive Rate | 0.11 | 0.12 |
| False Negative Rate | 0.55 | 0.68 |
| F1 Score | 0.19 | 0.19 |

The study found three significant predictors of HIV drug resistance in the surveyed populations. Notably, achieving HIV viral load suppression emerged as an important factor, with individuals who achieved suppression having a lower likelihood of developing drug resistance than those who did not. Furthermore, significant differences were found across countries, indicating potential disparities in healthcare practices or treatment accessibility. Additionally, adherence to antiretroviral therapy emerged as a critical factor, with individuals not on ARVs having a higher risk of drug resistance than those on ARVs.

**Figure 4:**
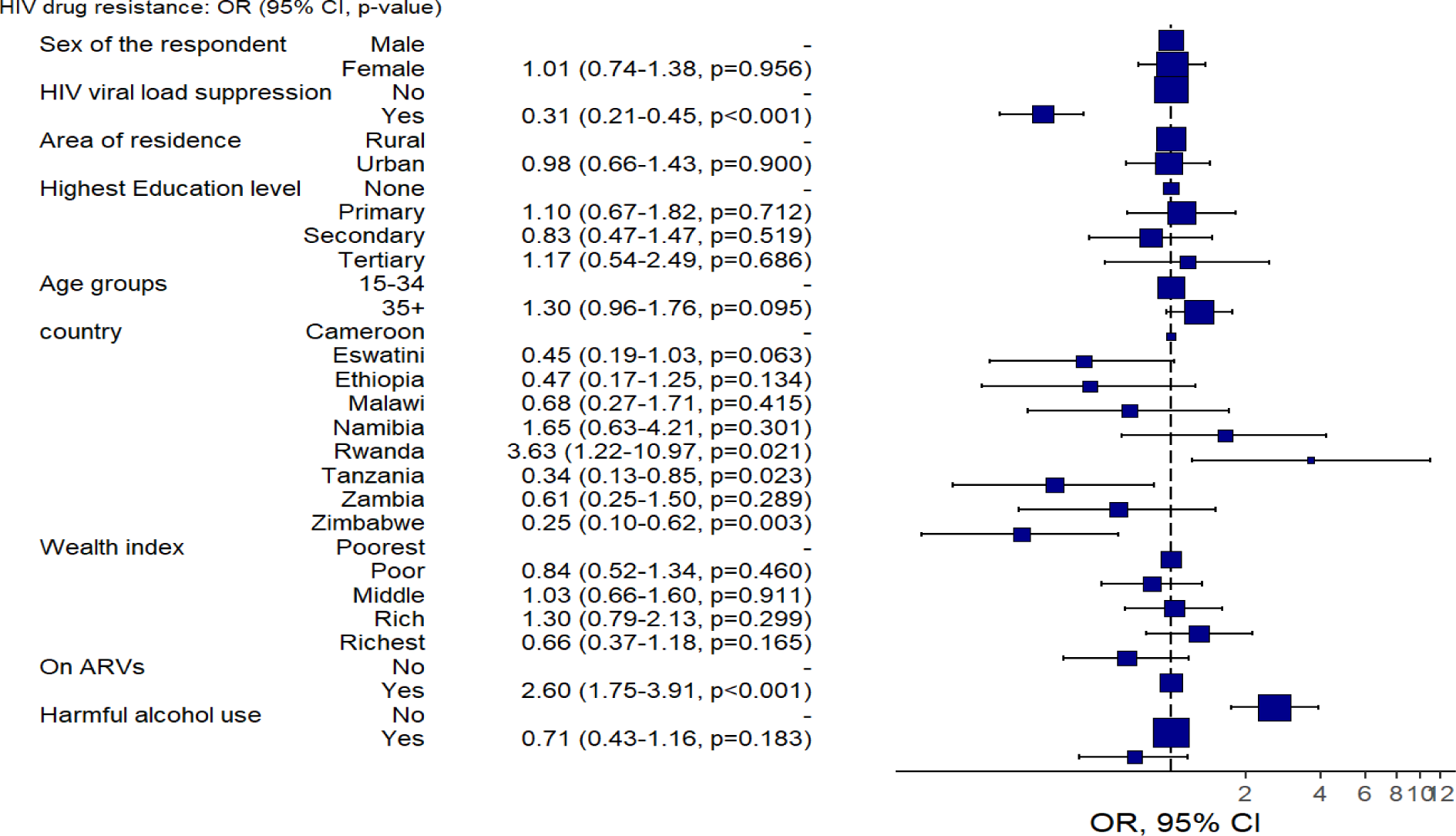
Predictive model of the outcomes using logistic regression and the forest plot of odds ratio of HIV Drug resistance in Rwanda, Namibia, Cameroon, Malawi, Eswatini, Zambia, Ethiopia, Zimbabwe and Tanzania between 2015 and 2019.

## DISCUSSIONS

The study highlighted the critical role of socioeconomic status (SES) in shaping patterns of HIV drug resistance across SSA. Lower-income individuals faced barriers to consistent treatment access and adherence, consistent with previous research. The association between unemployment rates and HIVDR rates underscores the intricate relationship between economic vulnerability and health outcomes, mirroring trends observed in other contexts(4).

In addition to socio-economic status (SES), demographic factors such as age and gender were identified as important predictors of HIV drug resistance. While lower-income individuals faced barriers to treatment access and adherence, the study also found that older age groups exhibited higher odds of developing drug resistance. This aligns with previous research indicating that age is a significant factor in HIV treatment outcomes, with older individuals experiencing greater challenges in adherence and response to therapy(8).

Moreover, gender disparities were observed in the study, with women constituting a majority of the population living with HIV in SSA. While gender did not show a significant association with HIVDR incidence, it is important to consider the unique challenges faced by women in accessing and adhering to HIV treatment. Sociocultural factors, such as gender norms and power dynamics, may influence treatment outcomes among women, underscoring the need for gender-sensitive interventions (7).

Furthermore, geographical disparities within SSA, particularly between urban and rural areas, were identified as contributing factors to the variability in HIVDR prevalence. Limited healthcare resources and disparities in access to care exacerbate challenges in effectively managing HIV drug resistance, echoing findings from studies conducted in other resource-limited settings (6).

Furthermore, the study highlighted the impact of clinical parameters on HIVDR, including viral load suppression. Individuals with unsuppressed viral load were found to be at increased risk of developing drug resistance. This suggested that disease progression and immune status play a crucial role in shaping the development of HIV drug resistance, emphasizing the importance of early diagnosis (5).

The study identified ARV status as a critical determinant of HIVDR, with individuals on antiretroviral therapy (ART) exhibiting varying levels of resistance compared to those not receiving treatment. Among individuals receiving ART, the prevalence of HIVDR was notably higher, indicating the emergence of resistance strains despite treatment efforts. This finding underscores the importance of treatment adherence, drug efficacy, and monitoring protocols in optimizing treatment outcomes and minimizing the development of drug resistance(9).

Furthermore, the study explored regional variations in HIVDR prevalence across different countries in SSA, highlighting disparities in healthcare infrastructure, treatment accessibility, and epidemiological contexts. This was in line with study by Moyo et al, where countries with limited healthcare resources and infrastructure were found to have higher rates of HIVDR. Moyo’s study further found that countries with robust healthcare systems and comprehensive ART programs demonstrated lower HIVDR prevalence, emphasizing the role of healthcare investments and policy frameworks in mitigating drug resistance(9).

The logistic regression analysis, performed on both training and test datasets in predictive modeling of HIVDR analysis using supervised machine learning provided critical insights on the key significant predictors of HIVDR. The implications of these results are significant for HIV/AIDS management and public health interventions in SSIA.

Firstly, the identification of key predictors of HIVDR, such as viral load suppression and antiretroviral therapy (ARV) status, country of residence suggests the need for tailored interventions. Healthcare providers can prioritize strategies aimed at achieving viral load suppression and promoting adherence to ARVs among individuals living with HIV/AIDS. Moreover, the observed variations in HIVDR across countries highlight the importance of addressing regional disparities in healthcare practices and treatment accessibility. Efforts to strengthen healthcare systems and improve access to HIV treatment and care should have priority, particularly in regions with higher rates of HIVDR.

Furthermore, Harmful alcohol users where more likely to have HIVDR than those who were none users, this is in line with the findings of the study done in Ethiopia, which found that alcohol abusers were more likely to develop HIVDR (19). This information is vital, as it will help decision makers to gain insight into the systems complexity to identify pointers for effective interventions. This methodology also applies when studying HIVDR in specific settings to gain insights into other complex problems. Moreover, the content of the model presented in this study applies to studies of other chronic diseases such as diabetes or obesity in order to understand their drivers and feedback loops (19).

The conceptual model presented here also lays the basis for quantitative mathematical modelling of the factors influencing HIVDR. This will allow quantitative modelers to collect data on relevant parameters in the system to monitor any changes, desired or not, in the entire system. An important advantage of basing a quantitative model on this conceptual map lies in the multidisciplinary manner in which the map was developed, therefore this helps in identifying mechanisms, which might not have been identified using a mono disciplinary approach (19).

Lastly, the application of predictive modeling techniques in identifying individuals at risk of HIVDR suggests the potential for integrating these approaches into routine clinical practice. Predictive models can aid healthcare providers in identifying individuals who may benefit from targeted interventions to prevent or manage drug resistance. Ultimately, these results can inform public health policy and resource allocation strategies aimed at addressing HIVDR in Sub-Saharan Africa, prioritizing interventions to reduce the burden of HIVDR and improve health outcomes for individuals living with HIV/AIDS.

Despite significant progress in HIV/AIDS research, prevention, and treatment, formidable challenges persist in our global fight against the epidemic. Drug resistance presents a formidable threat to the effectiveness of antiretroviral therapy, potentially undoing hard-fought progress and impeding efforts to curb HIV transmission. To surmount these challenges and advance towards the goal of ending the HIV/AIDS epidemic, comprehensive and coordinated action is imperative. This necessitates a holistic approach that combines biomedical interventions with initiatives addressing social determinants of health, inequalities, limited healthcare access, and underserved populations.

## AUTHORS’ CONTRIBUTIONS

EN and MG led the manuscript writing, WFN conducted data management and analysis; CZ advised on the data analysis and policy insights on the paper. All authors read and approved the final manuscript.

## Data Availability

The data used in this manuscript was derived from the Population-based HIV Impact Assessment Survey (PHIA). Please refer to the data access policies of PHIA for more information

https://phia-data.icap.columbia.edu/datasets

## ACKNOWLEDGEMENT

The authors would like to thank the ICAP at Columbia University for allowing us to use the data.

